# A scoping review of regulatory T cell dynamics in convalescent COVID-19 patients – Implications for Long COVID?

**DOI:** 10.1101/2022.10.04.22280642

**Authors:** Simon Haunhorst, Wilhelm Bloch, Florian Javelle, Karsten Krüger, Sabine Baumgart, Sebastian Drube, Christina Lemhöfer, Philipp Reuken, Andreas Stallmach, Michael Müller, Christina E. Zielinski, Mathias W. Pletz, Holger H.W. Gabriel, Christian Puta

**Author notes:** **Corresponding author:** PD Dr. Christian Puta, Department for Sports Medicine and Health Promotion, Friedrich-Schiller-Universität Jena, Wöllnitzer Straße 42, 07749 Jena, Germany.

## Abstract

**Background:** Recovery from coronavirus disease 2019 (COVID-19) can be impaired by the persistence of symptoms or new-onset health complications, commonly referred to as Long COVID. In a subset of patients, Long COVID is associated with immune system perturbations of unknown etiology, which could be related to compromised immunoregulatory mechanisms.

**Objective:** The aim of this scoping review was to investigate if regulatory T cell (Treg) dysregulation is observable beyond the acute illness and if it might be involved in Long COVID immunopathology.

**Design:** A systematic search of studies investigating Tregs during COVID-19 convalescence was conducted on MEDLINE (via Pubmed) and Web of Science.

**Results:** The literature search yielded 17 relevant studies, of which three included a distinct cohort of patients with Long COVID. The reviewed studies suggest that the Treg population of COVID-19 patients can reconstitute quantitatively and functionally during recovery. However, the comparison between recovered and seronegative controls revealed that an infection-induced dysregulation of the Treg compartment can be sustained for at least several months. The small number of studies investigating Tregs in Long COVID allowed no firm conclusions to be drawn about their involvement in the syndrome’s etiology. Yet, even almost one year post-infection Long COVID patients exhibit significantly altered proportions of Tregs within the CD4^+^ T cell population.

**Conclusions:** Persistent alterations in cell frequency in Long COVID patients indicate that Treg dysregulation might be linked to immune system-associated sequelae. Future studies should aim to address the association of Treg adaptations with different symptom clusters and blood parameters beyond the sole quantification of cell frequencies while adhering to consensualized phenotyping strategies.

## 1. Introduction

Infections with the severe acute respiratory syndrome coronavirus 2 (SARS-CoV-2), the pathogen responsible for the ongoing coronavirus disease 2019 (COVID-19) pandemic, manifest with varying severity. The clinical spectrum ranges from asymptomatic/mild disease to severe pneumonia and respiratory distress syndrome that can ultimately lead to death (1, 2). Although the majority of patients experience moderate symptoms such as fever, cough, dyspnea, loss of smell and taste or sore throat, it has been highlighted that COVID-19 can include multi-system complications, such as thrombotic events, vasculitis and myocarditis (3–5). In addition to that, the clinical spectrum of COVID-19 encompasses acute immunopathology. Specifically, dysfunctional cellular and humoral immune responses characterized by exaggerated cytokine release, lymphopenia and new onset or aggravated autoimmunity are associated with negative disease outcomes (1, 6–9).

It is estimated that approximately one in eight patients who contracted COVID-19 experience symptoms beyond the acute symptomatic phase, as reported by a recent observational cohort study (10). The World Health Organization (WHO) defines these sequelae, by common usage now also referred to as “Long COVID”, as a post-COVID-19 condition that usually occurs three months after a confirmed or probable SARS-CoV-2 infection with a set of new-onset, persistent or fluctuating symptoms that last for at least two months (11). The most common symptoms in this context are fatigue, exercise intolerance, post-exertional malaise, difficulty breathing, headache, muscle pain, tachycardia, concentration deficits and diminished quality of life (12–15). The pathophysiological mechanisms involved in the devolvement of Long COVID remain to be elucidated, with a range of theories currently under discussion. Merad et al. (16), for instance, postulated that immunopathological processes, such as chronic inflammation with viral persistence, post-viral autoimmunity, microbiome dysbiosis and unrepaired tissue damage might contribute to the pathophysiology of the Long COVID phenotype. In line with that, it has been documented that affected patients exhibit a significant elevation of multiple inflammatory markers compared to recovered subjects, indicating a dysregulated and overactive immune system (17, 18). Correspondingly, the triad of pro-inflammatory cytokines IL-1β, IL-6 and TNF-α showed a significant correlation with persistent symptoms of COVID-19 at an eight-months follow-up (19). In addition, the discovery of functional autoantibodies specific to G-protein coupled receptors (20) has been linked to an autonomic dysregulation and assumed to be a cause of residual symptoms (21).

Despite this, components of the immune system that regulate and fine-tune (auto)immune responses, favoring immune homeostasis, such as regulatory T cells (Tregs), have only been investigated sparsely in COVID-19 convalescence and Long COVID. Tregs are a dynamic CD4^+^ T cell subpopulation that is characterized by CD25 (IL-2 receptor α-chain) upregulation, CD127 (IL-7 receptor α-chain) downregulation and expression of the transcription factor forkhead box P3 (FoxP3) that is considered the master regulator of Treg development (22–25). It is estimated that 80% of the peripheral cell repertoire is represented by functionally mature natural Tregs that develop in the thymus as antigen-primed cells (26, 27), while peripherally-derived Tregs are induced from naïve T cells upon antigen encounter (27–29) (Figure 1). During self-directed and pathogen-directed immune responses, activated Tregs exert suppressive effects on effector cells and antigen-presenting cells, for example, through the secretion of the inhibitory cytokines IL-10, IL-35 and TGF-β, by limiting the amount of IL-2 available for conventional T cells or by stimulating the formation of indoleamine 2, 3-dioxygenase (23, 30) (Figure 1). In this way, Tregs promote self-tolerance and prevent excessive inflammation, which makes their proper functioning indispensable for a balanced immune response. Correspondingly, diseases such as type 1 diabetes, multiple sclerosis, lupus erythematosus and rheumatoid arthritis are associated with functional deficiencies in the Treg compartment and altered cell frequencies (31, 32).

**Figure 1:**
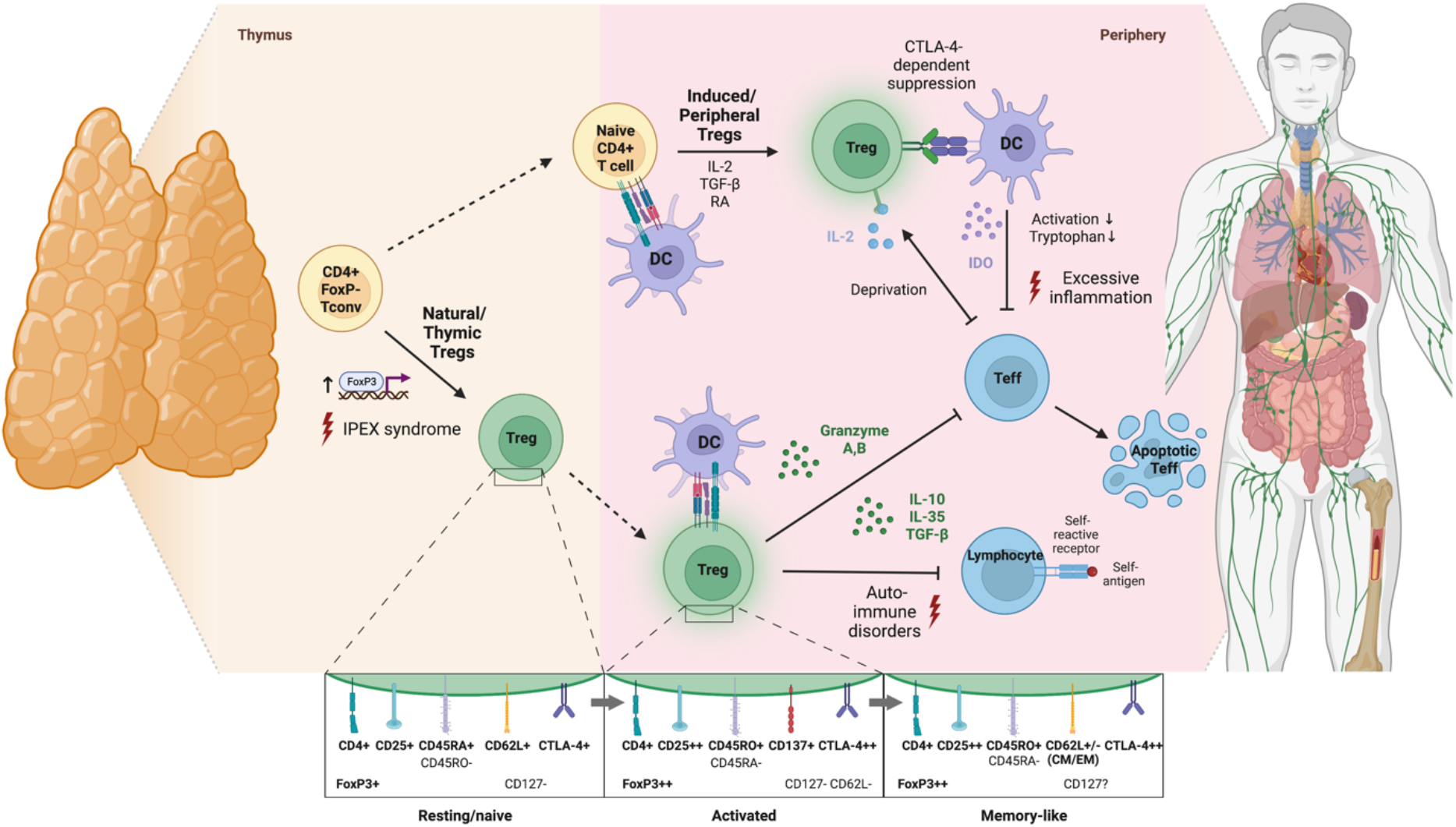
Schematic illustration of Treg development, modes of activation and physiological functions. According to an affinity-based model of T cell development, a fraction of self-reactive T cells with intermediate affinity for self-peptide-MHC complexes differentiates into natural Tregs by upregulation of FoxP3 and CD25 in response to T cell receptor (TCR) signaling (thymus-derived Tregs; ∼80% of peripheral Treg repertoire) (26). Also, antigenic stimulation of peripheral naïve T cells in the presence of TGF-b, IL-2 and retinoic acid induces their differentiation into FoxP3+ Tregs (peripherally-derived Tregs) (23). Of note, phenotypical transition of Tregs upon activation is not associated with de novo expression of surface proteins but rather with quantitative shifts in the expression levels of molecules already expressed in a resting state. Specifically, upon TCR activation, Tregs downregulate CD45RA and upregulate CD45RO, CD25, CTLA-4 and FoxP3 (27, 28). In contrast to other T cell subsets, the transition of Tregs into a stable memory pool after antigen elimination is still debated and so far, only a few marker candidates such as CD62L have been proposed for identification of memory Tregs (29). Functionally, Tregs are involved in suppressing immune responses towards self and non-self-antigens via inhibitory cytokine secretion, granzyme-dependent and cytokine-deprivation-mediated effector cell inhibition as well as cell-contact-dependent alteration of dendritic cell function and maturation (potential consequences of deficient Treg development or function are indicated by red lightnings) (30) (created with BioRender.com). *CM=central memory; CTLA-4=cytotoxic T lymphocyte antigen 4; DC=dendritic cell; EM=effector memory; IDO=indoleamine 2,3-dioxygenase; IPEX= Immunodysregulation, polyendocrinopathy, enteropathy, X-linked syndrome; RA=retinoic acid*

Studying COVID-19, previous reviews have already made an effort to characterize Tregs during the acute phase of a SARS-CoV-2-infection (33–36). Although evidence exists that alterations in the Treg compartment might contribute to the immunopathology of COVID-19, the literature concerning their functionality and frequency during the acute illness is controversial. Nevertheless, a trend toward functional impairments and decreased levels of circulating Tregs in severe cases of COVID-19 has been reported previously (33–35). It was shown that patients dying from COVID-19 exhibited significantly lower Treg counts and higher Th17/Treg ratio compared to recovered and healthy controls (37–39) and that low Treg counts at hospital admission were associated with clinical worsening and longer duration to discharge (40, 41). Correspondingly, several studies demonstrated a downregulation of FoxP3 in severe cases of COVID-19 (39, 42–44).

Given the aforementioned evidence suggesting that an ongoing inflammatory profile, autoantibody formation, inflammatory tissue damage, SARS-CoV-2 persistence and viral reactivation (herpes virus family) likely contribute to the persistence of symptoms, it is conceivable that Tregs are involved in the pathophysiology of Long COVID (45). Yet, there exist uncertainties concerning the longitudinal dynamics of the Treg population during recovery from COVID-19, and it remains unclear if Treg dysfunction is involved in the immunopathology of Long COVID. Therefore, we aimed to summarize the existing literature on the frequency (e.g., absolute counts, relative frequency) and functionality (e.g., cytokine secretion, ex vivo suppressive capacity) of Tregs in convalescent COVID-19 patients with and without persisting symptoms.

### 2. Methods

A scoping review of the literature was conducted following the Preferred Reporting Items for Systematic Reviews and Meta-Analysis (PRISMA) extension for scoping reviews (46).

### 2.1 Literature search

The literature search was conducted up to July 23, 2022, using Pubmed and Web of Science with no restriction on publication date. The search string included controlled vocabulary, Medical Subject Headings (MeSH) and common synonyms for the domains COVID-19 and regulatory T cells while excluding review articles (see appendix). Furthermore, reference lists of included studies were examined in order avoid missing any relevant studies.

The titles and abstracts of the records that were identified through the literature search were independently screened for their eligibility by two investigators (SH and CP). Subsequently, full-text articles were retrieved to verify the inclusion decision that was made based on title and abstract screening (Figure 2).

**Figure 2:**
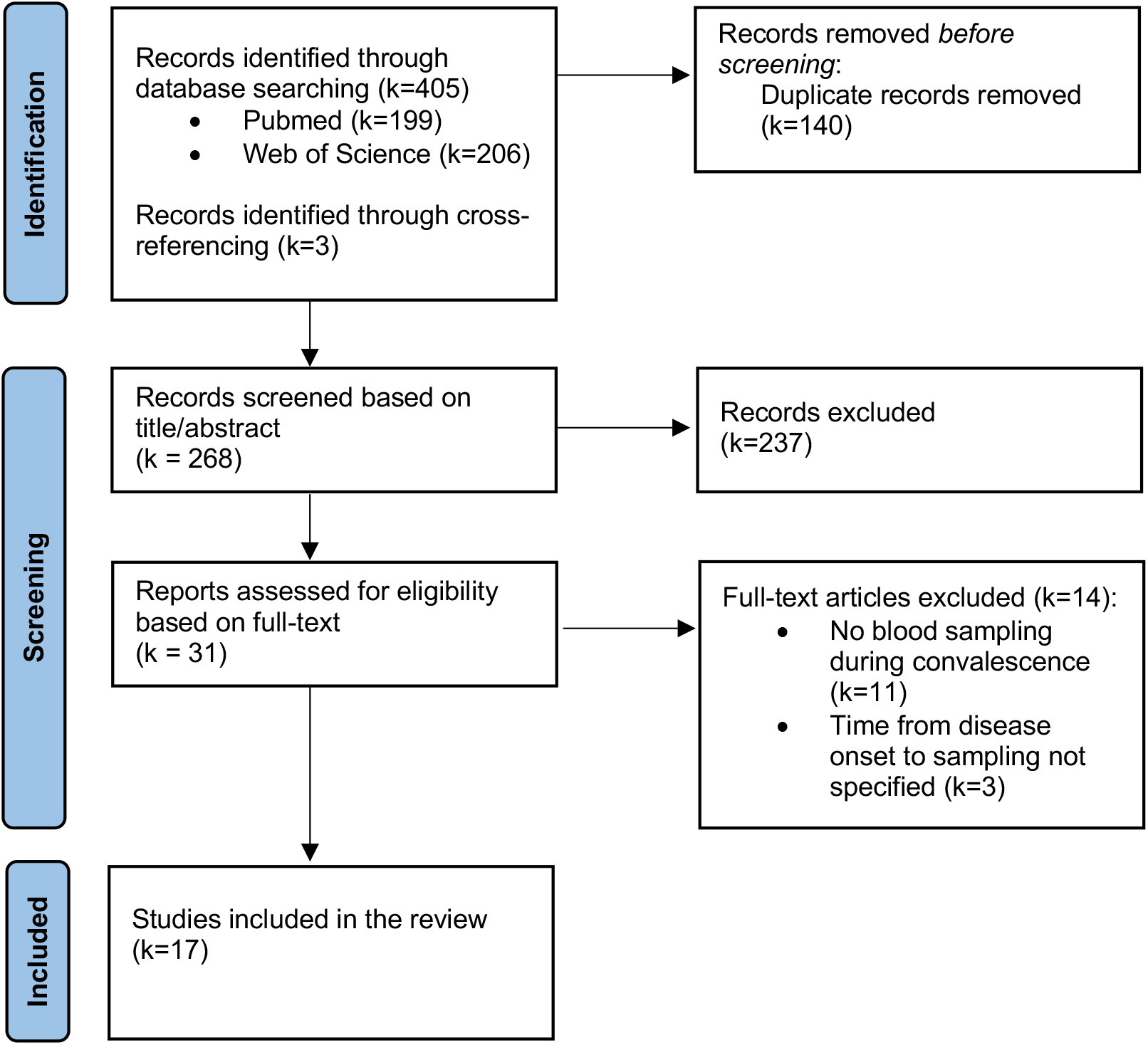
Flow diagram illustrating the literature search and study selection.

### 2.2 Study selection and eligibility criteria

The eligibility criteria were formed a priori using the PICOS (participants, intervention, comparators, outcomes, and study design) approach. We considered studies to be eligible for inclusion in this review if they met the following criteria: (1) population: a cohort of patients at least four weeks after confirmed or probable detection of infection with SARS-CoV-2; (2) intervention: mass or flow cytometric analysis of Treg outcomes; (3) outcomes: Treg counts, proportions or regulatory function, or FoxP3 gene expression; (4) study: case-control studies, cross-sectional studies, cohort studies, clinical trials, or case reports published in English or German. Book chapters, congress proceedings, meeting abstracts, reviews and studies published in languages other than English or German were deemed ineligible.

### 2.3 Data extraction and synthesis

The following data items were extracted from the included studies: sample size, sex, gender, time elapsed since disease onset and severity of acute disease, markers used for Treg phenotyping, source of the cell sample, Treg-related outcome measures and outcome-related findings. Extracted data items are presented in a tabulated form (Table 1).

**Table 1.**
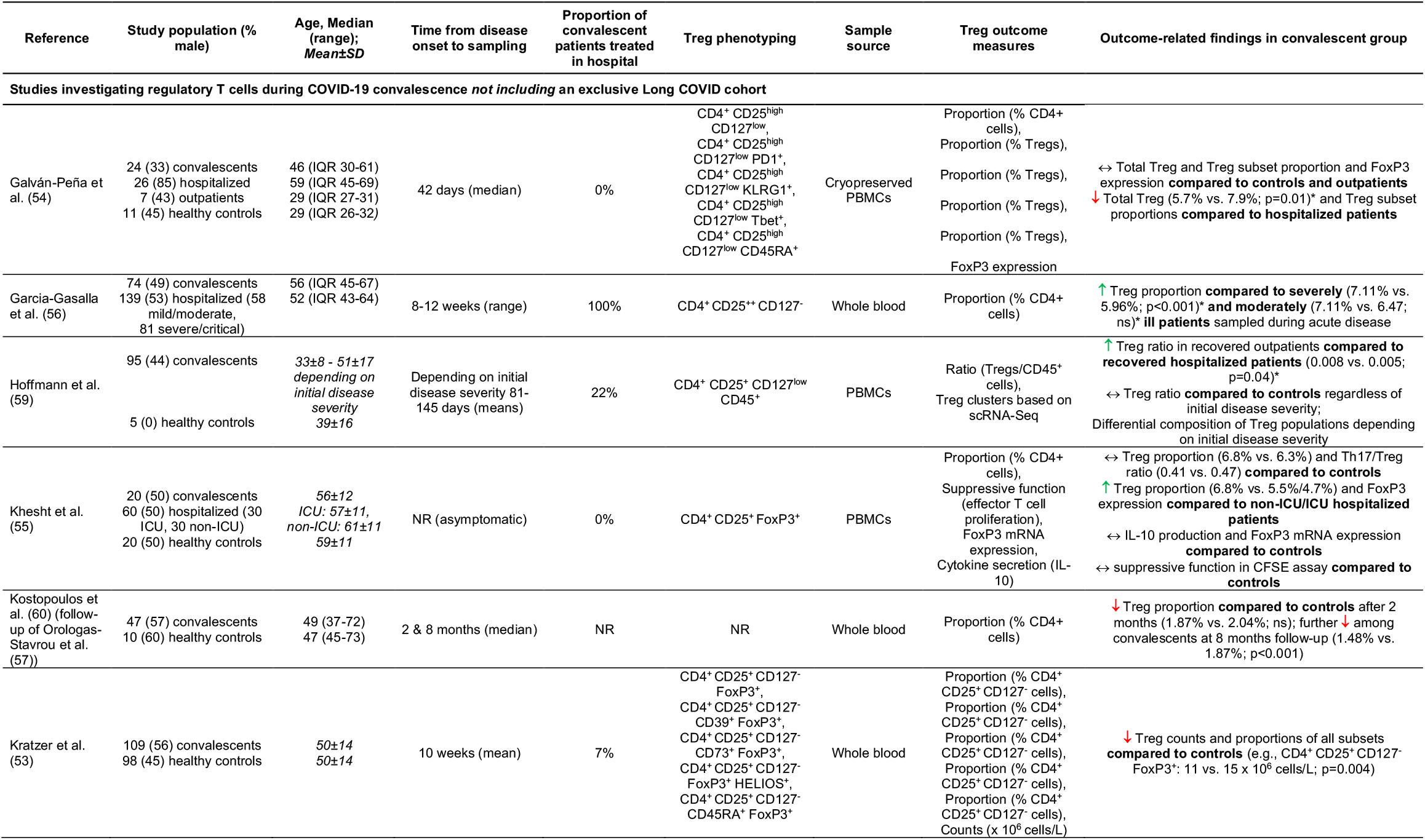
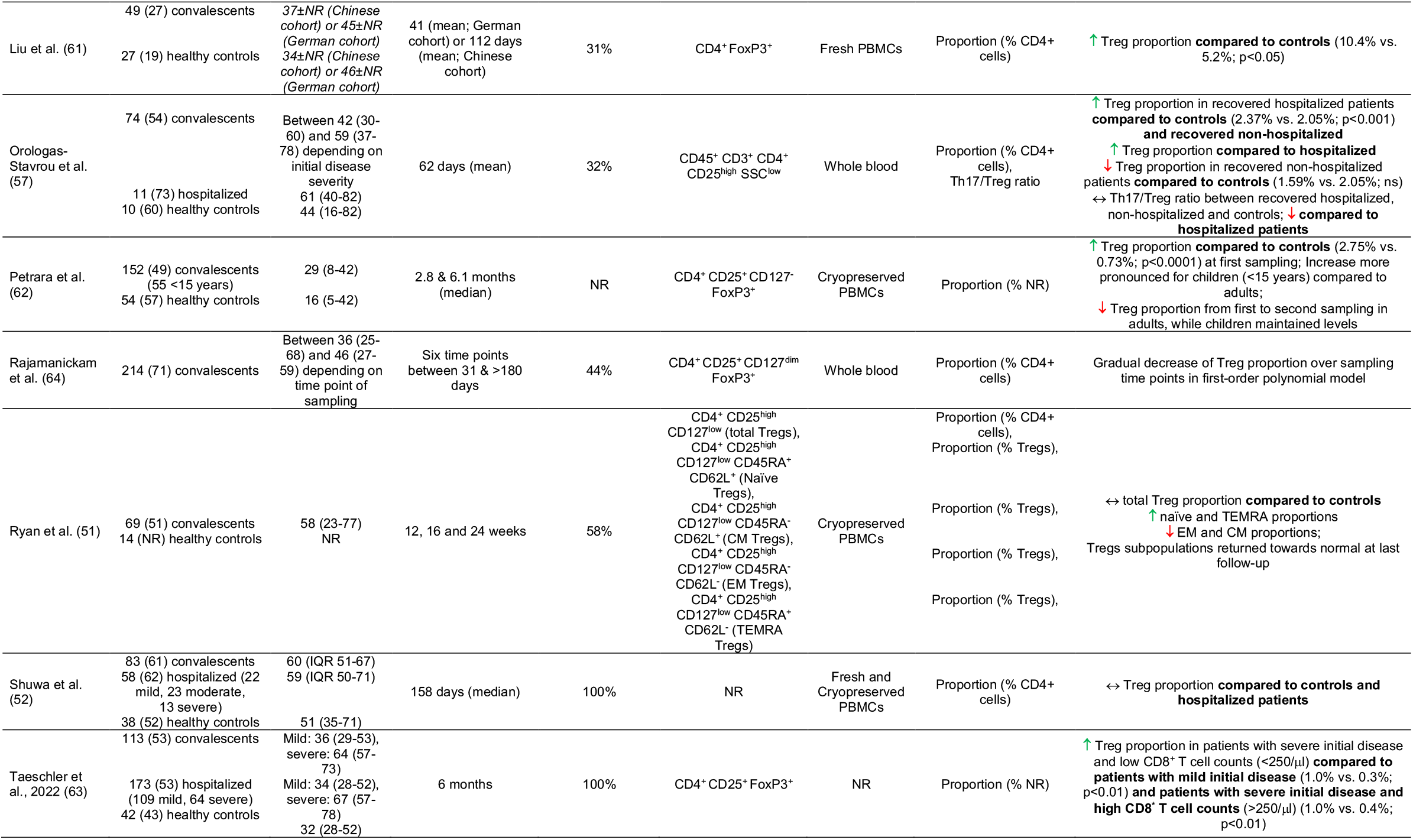
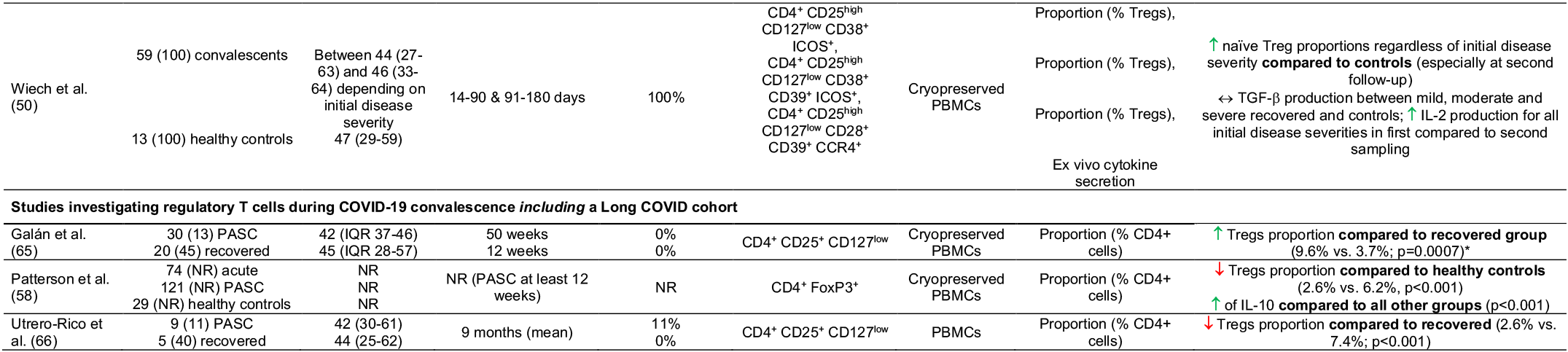
Characteristics and main results of the included studies. *CM Tregs=central memory Tregs; CFSE= Carboxyfluorescein succinimidyl ester; EM Tregs=effector memory Tregs; ICU=intensive care unit; IQR=interquartile range; NR=not reported; ns=not significant; PASC=post-acute sequelae of COVID-19 infection; PBMCs=peripheral blood mononuclear cells; scRNA-Seq=single cell RNA sequencing; SD=standard deviation. * Extracted from plotted data using WebPlotDigitizer software (https://automeris.io/WebPlotDigitizer/)*

## 3. Results

The literature search yielded a total of 405 potentially relevant records. After removing duplicates, 268 studies were screened for their eligibility, of which 17 were ultimately included in this review. Three studies (47–49) were deemed potentially relevant but could not be included due to insufficient characterization of the convalescent cohort in terms of the time point of blood sampling (Figure 2).

### 3.1 Study characteristics and investigated outcomes

All studies investigated a cohort of patients with a confirmed or presumed COVID-19 infection at least four weeks and latest 50 weeks after index infection. The total number of participants investigated during COVID-19 convalescence was 1367 (range: 14-214). In addition, 13 studies included a cohort of seronegative controls (371 total participants), and seven included one or more cohorts of patients sampled during acute COVID-19 illness (375 total participants of which 368 had been admitted to the hospital). The number of studies characterizing Tregs in a cohort of patients with Long COVID, in accordance with the previously mentioned definition by the WHO, was limited to three, with a total number of 160 participants (Table 1).

Eleven of the included studies did not recruit an exclusive cohort of patients specifically diagnosed with Long COVID, thus we must assume that the convalescent patients investigated in these studies were all recovered from COVID-19 and did not exhibit any residual symptoms. Wiech et al. (50) assessed Long COVID symptoms of investigated subjects retrospectively at a one-year follow-up and reported that 72% of the participants experienced persistent symptoms (e.g., cognitive dysfunction, fatigue, dyspnea) for at least three months. Likewise, Ryan et al. (51) retrospectively assessed that 30% of the participants were referred to a Long COVID clinic and Shuwa et al. (52) reported that approximately half of the patients experienced dyspnea at follow-up sampling. However, since those subjects with persisting symptoms were analyzed together with recovered subjects, the results of these studies provide only limited evidence concerning Treg outcomes in Long COVID.

All studies measured the relative cell frequency as the central Treg-related outcome, whereas only one study reported absolute cell counts (53). In addition to that, the expression level of the FoxP3 transcription factor was investigated in two studies (54, 55) and domains of Treg functioning like suppressive function and cytokine production in one (55) and two (50, 55) studies, respectively. Studies analyzed whole blood or peripheral blood mononuclear cells using flow cytometry and mass cytometry. The markers used for Treg phenotyping differed markedly between the studies and are detailed in Table 1 together with the study and subject characteristics.

### 3.2 Tregs during COVID-19 convalescence

#### 3.2.1 In comparison to acutely ill patients

Although no study compared blood samples obtained from the same cohort of patients both during acute illness and convalescence, six studies compared Treg parameters between recovered and acute cases of different severity (Figure 3). A significant difference in peripheral Treg frequency in relation to recovering patients could only be demonstrated for acutely hospitalized patients, whereas non-hospitalized patients exhibited no different Treg proportions. Garcia-Gasalla et al. (56) investigated 74 patients between eight and 12 weeks after hospital discharge and compared their immune profiles to patients that were sampled within the first 48 hours after hospital admission with different disease severity. Accordingly, the recovered patients exhibited greater relative Treg frequencies compared to severe/critical (p<0.001) and mild/moderate (not significant) cases that were hospitalized for COVID-19 (56). This is in line with other studies that demonstrated an expansion of the Treg population and also a decreased Th17/Treg ratio compared to severely ill patients, albeit the investigated convalescent patients mostly recovered from mild or asymptomatic diseases (55, 57). On the other hand, only one study documented a decrease in the Treg frequency in convalescent compared to hospitalized COVID-19 patients at a median follow-up of 42 days after mild infection (p=0.01) (54). This change could however not be observed in comparison to outpatients (54). Likewise, another study found no change between patients recovering from severe COVID-19 and acutely ill patients, regardless of their disease severity (52).

**Figure 3:**
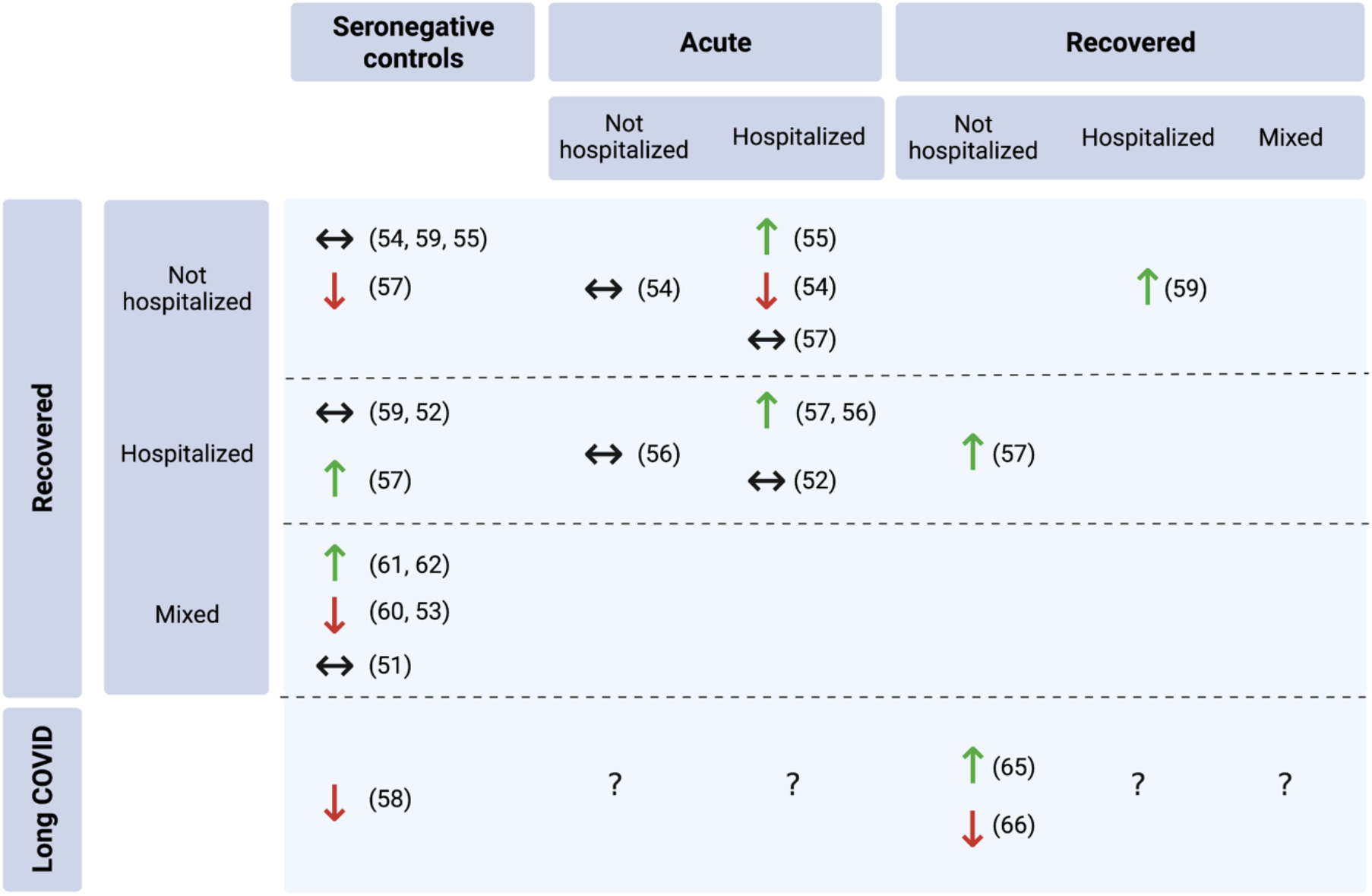
Total Treg frequency in convalescent COVID-19 patients compared to other cohorts (references in brackets). The upwards arrow (τ) indicates a significant increase at p<0.05, the downwards arrow (τ) a significant decrease at p<0.05 and the left-right arrow (ττ) not significantly different relative Treg proportions in convalescent patients (created with BioRender.com).

Adding to this body of literature, several studies assessed parameters of Treg functionality in relation to acute cases. Specifically, Galván-Peña et al. (54) reported that Tregs from severe acute cases showed overlaps with tumor-infiltrating Tregs. Specifically, the analysis of the transcriptional signature revealed for example an acute enrichment of hypoxia-induced transcripts that was not present in healthy donors and only scarcely in recovered patients. Furthermore, the expression of FoxP3, IL-10 production and the suppressive function on effector T cell proliferation has been reported to be higher in recovered compared to severely ill patients (55, 58). However, it bears noting that all patients that were investigated for Treg functioning during convalescence recovered from mild disease.

#### 3.2.2 In comparison to seronegative controls

The comparison between recovered patients and seronegative controls produced ambiguous results. While five studies found no significant difference in frequencies of total Tregs between patients post-COVID-19 and patients without history of COVID-19 (51, 52, 54, 55, 59), two studies reported a lower (53, 60) and four reported a greater peripheral Treg frequency in recovered patients (50, 57, 61, 62) (Figure 3). However, this discrepancy did not seem to be attributable to differences in initial disease severity or the time elapsed since diagnosis. A study of 49 individuals recovering from COVID-19 in two different countries, of which approximately one-third were hospitalized, showed that the COVID-19 group exhibited significantly higher Treg frequencies than healthy controls at a mean follow-up of 41 or 112 days (p<0.05) (61). In line with that, Petrara et al. (62) demonstrated greater Treg proportions in patients several months after index infection compared to uninfected controls (p<0.0001), with that difference being even greater in the pediatric (<15 years) population. By contrast, Kratzer et al. (53) reported lower total Treg cell frequencies in individuals recovering from predominantly mild infections (p=0.004), which was further accompanied by lower proportions of several Treg subsets compared to controls (p<0.006).

A more detailed analysis of Treg subsets in a cohort of convalescent patients that did not exhibit significantly different total Treg proportions compared to controls showed that subsets in different maturation statuses differed significantly between investigated groups. Accordingly, naïve Tregs (CD4^+^ CD25^high^ CD127^low^ CD45RA^+^ CD62L^+^) and effector memory Tregs expressing CD45RA (TEMRA; CD4^+^ CD25^high^ CD127^low^ CD45RA^+^ CD62L^-^) were more frequent, whereas central memory (CD4^+^ CD25^high^CD127^low^CD45RA^-^ CD62L^+^) and effector memory (CD4^+^ CD25^high^ CD127^low^ CD45RA^-^ CD62L^-^)

Tregs were lower in patients 12 weeks post-infection (51). Consistent with this, Wiech et al. (50) documented increased proportions of naïve cells among Tregs compared to healthy donors.

One study assessing the cytokine release of Tregs in seronegative controls reported that there was no significant difference in IL-10, IL-17 production and suppressive function compared to previously asymptomatic patients (55).

#### 3.2.3 Comparisons between convalescent patients recovering from diseases with different severity

Five studies compared Treg outcomes among recovered patients with different initial disease severities, reaching equivocal results (50, 51, 57, 59, 63). A study that followed-up subjects that were hospitalized for mild and severe courses of COVID-19 found out that six months post-discharge there were patients with severe illness that presented with persistently low CD8^+^ T cell counts (<250/ml) (63). Those subjects exhibited a higher Treg frequency than severely ill who were categorized as having high CD8^+^ T cell counts (>250/ml) and patients with mild initial disease (63). Two other studies that investigated total Treg frequencies documented both higher and lower proportions in convalescent outpatients compared to those who were hospitalized (51, 59) (Figure 3). Similarly as heterogeneous were the results of the studies that investigated Treg subsets. While Ryan et al. (51) found no considerable difference between patients recovering from mild and severe initial disease for naïve, central memory, effector memory or TEMRA subsets, another study reported lower effector Treg populations and lower FoxP3 expression levels in previously hospitalized patients (59). At the same time, single-cell RNA sequencing analysis revealed that relative to all other severity groups, patients recovering from an asymptomatic infection exhibited an expansion of the Treg population with high expression of class II HLA molecules that are suggested to exert high contact-mediated suppression of T helper cell responses (59).

#### 3.2.4 Longitudinal assessment in convalescent patients

Six studies obtained blood samples at multiple time points during COVID-19 convalescence (50, 51, 59, 60, 62, 64). The longitudinal analysis conducted by Kostopoulos et al. (60) showed that the relative frequency of total Tregs that was decreased two months post-infection continued to decrease at the eight-month follow-up (p<0.001). Similarly, Wiech et al. (50) reported that naïve Tregs further increased from first (14-90 days post-infection) to second (91-180 days post-infection) follow-up regardless of the patients’ initial disease severity, whereas central memory Tregs decreased. Additionally, the in vitro production of IL-2 decreased significantly over time, while the TGF-β production did not change (50). Another three studies demonstrated that Treg frequency and parameters of Treg functioning that were up-or downregulated earlier in the recovery period, returned to an equilibrium point (51, 59, 62). Accordingly, Ryan et al. (51) documented that naïve and TEMRA subsets that were greater as well as effector memory and central memory subsets that were lower in recovered subjects 12 weeks post-infection, decreased or increased towards the values exhibited by seronegative controls at the 16- and 24-weeks follow-ups.

#### 3.2.5 Tregs in Long COVID

Three studies investigated the proportion of Tregs among CD4+ cells in patients that exhibit residual symptoms of COVID-19 and compared them to seronegative controls and individuals that recovered from COVID-19 (58, 65, 66) (Figure 3). The symptoms experienced by the patients in the Long COVID group covered a broad spectrum, including concentration and memory impairments, headaches, palpitations, insomnia, myalgia, fatigue and shortness of breath. The two studies that sampled patients with lingering symptoms almost one year post-infection reported contradictory results. While Galán et al. (65) documented a 2.5-fold greater Treg frequency in Long COVID patients compared to subjects that recovered completely (p=0.0007), Utrero-Rico et al. (66) found a change in the opposite direction (p<0.001). Likewise, 121 Long COVID patients exhibited significantly lower proportions of FoxP3-expressing CD4+ cells compared to seronegative controls (58). None of the studies investigated parameters of Treg functioning such as cytokine secretion or suppressive capacity in Long COVID.

## Discussion

The objective of this scoping review was to summarize the existing literature regarding the frequency and functionality of Tregs in convalescent COVID-19 patients and to explore indications for their potential involvement in the development of Long COVID. Owing to the fact that none of the studies investigating a cohort of convalescent COVID-19 patients obtained baseline blood samples during the acute phase of the disease, the exact longitudinal dynamics of Tregs after infection with SARS-CoV-2 remain elusive. However, the comparisons of separate cohorts of patients sampled during severe acute disease and recovery showed increased systemic frequency of Tregs and greater suppressive function in recovering subjects, which points towards a possible restoration of Treg frequency and function (55–57).

Earlier studies investigating patients during acute illness revealed that COVID-19 can induce alterations in lymphocyte frequency and functioning that are associated with disease severity and prognosis (2, 40, 67–69). In severe cases, a loss of CD4^+^ lymphocytes, including Tregs is a prominent clinical feature that, together with an enhanced myelopoiesis, contributes to an expansion of neutrophils, dendritic cells and macrophages, favoring immunopathology, excessive tissue infiltration and acute respiratory distress syndrome (40, 41, 70–73). Of note, the decrease of Tregs in severe COVID-19 patients is not only a reflection of diminished absolute cell counts in the context of lymphopenia but is also linked to altered relative cell frequencies, as a result of Th17 cell expansion (74).

The trend towards a quantitative reconstitution of Tregs was supported by five studies investigating patients recovering from diseases of different severity (51, 52, 54, 55, 59). The fact that the studies were not able to find significant differences in Treg frequency between recovering subjects and seronegative controls suggests that the peripheral Treg population was either unaffected by the initial disease or returned to normal values within the first six weeks following mild disease (54, 55, 59) or four months following severe disease (51, 59). Further illustrating this, three studies that obtained multiple blood samples during recovery from COVID-19 showed that the Treg frequency exhibited by subjects at the second and third follow-ups was closer to the frequency of seronegative controls than during the first follow-up (50, 51, 62). Beyond that, the expression of the Treg-defining transcription factor FoxP3 has been described to be significantly higher in convalescent patients (55). Yet, the significance of this finding is unclear, as the recruited subjects recovered from asymptomatic infections, which might induce less pronounced Treg perturbations in the first place, as it has been indicated by previous reviews (33, 35). Correspondingly, in comparison to acute non-hospitalized patients, recovered patients did not show significantly different Treg levels (52, 54, 56).

Nevertheless, it also became apparent that infection-induced adaptations in the Treg compartment are heterogeneous and that Treg dysregulation can persist for at least several months post-infection. Correspondingly, several of the reviewed studies found that compared to seronegative controls, recovering subjects exhibited significantly different Treg frequencies (53, 57, 60–62). Kostopoulos et al. (60), for example, reported that even eight months after disease onset the proportion of Tregs had not returned to normal. Beyond that, Shuwa et al.’s (52) findings showed that the range of the Treg frequency, as well as the deviation from the mean within the recovering group was considerably greater than what would be expected based on the data from seronegative controls, which indicates that SARS-CoV-2 might induce Treg adaptations that are highly individual and dependent on factors like age, gender, co-morbidities, training status or immunological homeostasis and epigenetic landscape (75–77).

Still, the studies reviewed here did not allow to draw any firm conclusions on how alterations in Treg frequency or function are associated with initial disease severity and time since disease onset. It is therefore yet unsolved what the mechanisms behind persistent dysregulations of the Treg compartment are and what factors determine if the immunological homeostasis is reinstated. Several mechanisms have been proposed to which the acute reduction of Tregs, especially in severely ill patients, might be attributable. The surge of IL-6 caused by an exaggerated innate immune response for example might selectively antagonize the generation of Tregs and FoxP3 induction while promoting a pro-inflammatory Th17 cell profile (78, 79). Supporting this point, a positive correlation between IL-6 and Th17, a greater Th17/Treg ratio, as well as an inverse correlation between neutrophil and lymphocyte numbers have been documented in severe cases of COVID-19 (80). Additionally, other factors that are important for Treg development, such as dehydroepiandrosterone sulfate, which inhibits IL-6 and activates Tregs as well as retinoic acid, which is essential for Treg differentiation are depleted in patients with COVID-19 (81, 82). Finally, Treg extravasation and compartmentalization into inflamed tissues and an increased apoptosis rate could be responsible for low peripheral Treg levels in a subset of patients (35).

The scarcity of studies investigating Tregs in this specific cohort allows no firm conclusions to be drawn about the nature of Treg adaptations in Long COVID. Still, the reviewed studies indicated that Long COVID patients exhibit Treg dysregulations long after their initial infection with SARS-CoV-2. Two studies that were comparable in terms of the time elapsed from disease onset to follow-up sampling, markers used for cell phenotyping and the subjects investigated, found both a significantly greater and lower proportion of Tregs among CD4+ cells in patients with residual symptoms compared to recovered subjects (65, 66). A decreased Treg frequency compared to seronegative controls was reported in a study of 121 subjects with post-acute sequelae of COVID-19 (58).

When interpreting these results, it must be kept in mind that Long COVID constitutes a heterogenous and multifaceted syndrome, presenting diverse clinical manifestations (83, 84). Previous research has proposed that several phenotypes and distinct sub-diagnoses might exist under the umbrella of Long COVID, some of which with little immunological contribution (85–87). Since the post-acute sequelae of COVID-19 can most certainly not be attributed to a single pathophysiological mechanism, it is conceivable that a Treg dysregulation contributes to a Long COVID-associated immunopathology in multiple ways (Figure 4), as it has also been suggested in other viral infections. For instance, the investigation of patients infected with hepatitis C virus (HCV) showed that those with chronic infections and HCV persistence exhibited higher Treg levels than recovered subjects (88, 89). By contrast, chronically infected patients that developed autoimmune complications exhibited significant reductions in Treg levels compared to those who did not and to healthy controls (89, 90).

**Figure 4:**
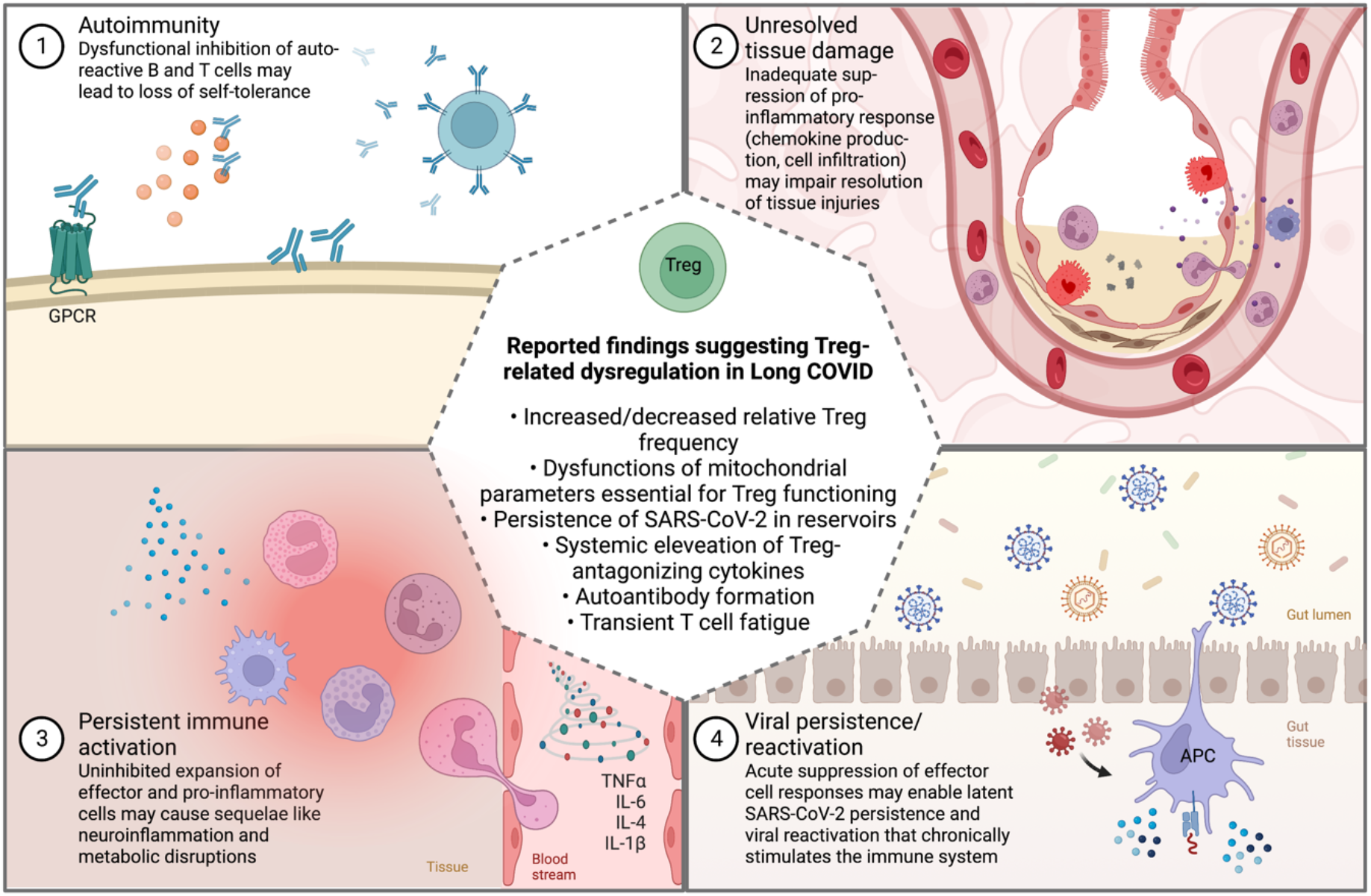
Commonly suggested theories for the non-exclusive pathophysiological mechanisms involved in Long COVID (1-4) and how they might be related to Treg dysregulation (created with BioRender.com). *GPCR=G-protein coupled receptor*

Post-viral autoimmunity is also one of the main hypotheses discussed with regard to the development of Long COVID (Figure 4). Functional autoantibodies against a wide array of cell components and messenger molecules have been identified in patients with persistent symptoms (20, 91–93). Clinically, the formation of vasoactive autoantibodies directed against G-protein-coupled receptors (GPCR), for example, might consequently impede vascular tone and autonomic functions. Correspondingly, the treatment with a DNA aptamer drug neutralizing autoantibodies targeting GPCR improved retinal capillary microcirculation and led to the disappearance of Long COVID symptoms (94). If Long COVID patients with latent autoimmunity also exhibit low levels or dysfunctional Tregs warrants further investigation.

On the other hand, an increase in the number and function Tregs can result in a suppression of effector immune responses, ultimately favoring deficient viral clearance and persistent infections that cause cyclical symptoms flare-ups (89) (Figure 4). The persistence of SARS-CoV-2 RNA and infected cells in certain tissue reservoirs, such as the olfactory mucosa or the gastrointestinal tract is a common observation that seems to be facilitated by immunosuppression (95–98). Persistent detection of viral RNA in plasma, stool and cerebrospinal fluid has further been reported in patients with Long COVID (99–101). Whether viral persistence in these patients is a reflection of perturbed Treg dynamics or rather the virus’ own capability to evade immune responses and to shield its RNA from degradation remains to be delineated. A study that examined molecular features of 50 patients with long and short duration of viral shedding found that Treg frequency was significantly higher in those with long duration shedding, accompanied by a general immunosuppressive status (102). Likewise, other studies demonstrated that the total Treg frequency was associated with prolonged SARS-CoV-2 positivity and RNA shedding (103, 104), collectively suggesting that high Treg levels constitute one factor that might favor viral persistence. Furthermore, Taeschler et al. (63) provided evidence that persistently low levels of CD8^+^ T cells six months post-discharge were associated with a higher Treg frequency.

However, it bears noting that an increased Treg frequency is not necessarily a reflection of a greater regulatory capacity within the cell population (25). The studies reviewed here did not specifically investigate aspects of Treg function in Long COVID, but some reports raise the possibility that pathophysiological mechanisms that are associated with Long COVID could also adversely affect Treg functions. It has, for example, been shown that the integrity of specific intracellular metabolic processes is essential for the maintenance of Treg suppressive function and that the reliance on mitochondrial metabolism is greater relative to other cells of the CD4 lineage (105, 106). Evidence indicating a mitochondrial dysfunction in Long COVID that compromises this metabolic integrity comes from studies that reported a loss of mitochondrial membrane potential (107), greater glycolysis rates (108, 109) or oxidative stress (110)in Long COVID. Additionally, persistently high systemic and tissue-level concentrations of inflammatory cytokines like IL-6 and TNF could subvert the Tregs’ regulatory capacity (111). For future studies, it would therefore be desirable to get further in-depth insights into Treg functional aspects in Long COVID and COVID-19 convalescence in general. In this context, it might also be important to consider the migratory capacity of Tregs, since the selective expression of adhesion molecules and chemokine receptors is a major determinant of their regulatory function (112). As Tregs play a decisive role in resolving tissue injuries (113, 114), it might also be worthwhile to investigate them in tissue samples from sites in which persistent inflammation and damage to the organ infrastructure have been documented throughout the post-acute phase, such as the lungs, myocardium or intestines (115–118).

Owing to the relative novelty of the research area, the studies reviewed in this scoping review had several limitations that need to be considered when interpreting their results, which could be harnessed to improve future research efforts in this area. A general issue that can limit the comparability of study results is the identification and isolation of Tregs. An extensive list of potential Treg marker candidates has been proposed (119). However, the fact that Tregs represent a highly dynamic cell population and that none of the markers is exclusively expressed by Tregs makes their characterization a complicated task (30). To facilitate standardized and reliable isolation procedures across studies, Santegoets et al. (120) published a consensus on essential markers for the detection and functional analysis of Tegs. They concluded that basal phenotyping should minimally include staining for CD4, CD25, CD127 and FoxP3, a recommendation that only three studies (53, 62, 64) sufficiently adhered to. Moreover, the studies included here used five different marker sets to identify total Treg frequencies, which might account for the broad range of Treg proportions across studies that was also present among seronegative controls.

None of the studies reviewed here explicitly stated which SARS-CoV-2 variant the included subjects contracted. Twelve studies conducted participant recruitment before the alpha variant was designated a variant of concern in December 2020. Therefore, it can be assumed that participants in these studies contracted the wild type. Four studies did not specify the date of sampling (54, 55, 58, 59)and Wiech et al. (50) reported that during sampling in early 2021 the alpha variant became dominant in Poland. Given that participant recruitment was carried out relatively early after the global emergence of SARS-CoV-2, it is also likely that most subjects have only been infected once. Furthermore, as the widespread rollout of vaccination campaigns in Europe and the United States began in early 2021, subjects investigated in the included studies were most certainly unvaccinated. While this homogeneity between studies in terms of the stage of viral evolution and lack of population immunity at the time of infection might enhance the comparability of results, it also leads to the circumstance that the presented data provide a temporary snapshot that is not necessarily representative of the current clinical situation. Over the last two years, several new variants have emerged that can effectively evade immunity acquired through vaccination or natural infection (121). The latest omicron variant encodes 37 amino acid substitutions in the spike protein, 15 of which are in the receptor binding domain, resulting in an antigenic variation that subverts innate and adaptive immunity and enhances cell entry (122, 123). Besides that, previous research has also indicated that many pathogens have developed mechanisms to use immune regulatory networks for their advantage, for example by upregulating the expression of IL-10 or TGF-beta and by dampening effector cell responses (89). Preliminary evidence showed that in vitro stimulation with omicron variant peptide induced significantly greater FoxP3 expression in draining lymph node cells than with the wild-type, which was interpreted as an indication that the variant peptide might induce a switch in T cell function from an effector to a regulatory program (124). In this context, it is also important to mention that none of the included studies investigated Tregs with regard to their antigen specificity. Taking these considerations into account, it should be further investigated if Treg adaptations are affected by the variant contracted and if they are altered in individuals that already had some level of SARS-CoV-2 specific immunity. Furthermore, the investigation of SARS-CoV-2-specific Tregs would provide deeper insights into the virally induced landscape.

Beyond that, it becomes more and more apparent that Long COVID cannot be regarded as a single diagnosis but that it encompasses several diagnoses with distinct pathophysiological mechanisms. It can therefore be assumed that given the range of symptoms exhibited by the included patients, the three studies investigating Long COVID portrayed a mix of different pathologies. For future studies, it would be intriguing to investigate if Treg frequencies and Th17/Treg ratio correlate with specific symptom clusters like they were previously proposed or with laboratory parameters such as serological status, cytokine levels or (auto)antibody titers.

## 5. Conclusions and perspectives

COVID-19 is associated with perturbations in the Treg homeostasis, albeit reports about exact changes in cell number remain controversial. The studies reviewed here indicated that dysregulation in the Treg compartment can persist for at least several months post-infection. Also, the results of the three studies included in this review, accompanied by previously postulated hypotheses regarding the development of Long COVID suggest that Treg dysregulation might be involved in the post-acute persistence of symptoms. However, the methodological heterogeneity of the included studies with respect to the subjects recruited, the phenotyping strategies and the sampling at different stages of the recovery makes it difficult to delineate what factors determine the reinstatement of immunological homeostasis. Collectively, it became apparent that despite plausible connections to the pathophysiology of COVID-19 and its associated sequelae, to this day, Tregs have received little scientific attention that goes beyond the quantification of cell numbers. Correspondingly, several aspects, such as alterations in Treg functionality need further investigation. Others like the influence of new variants, vaccination status and previous infections remained completely unaddressed in the included studies and should be considered for future research efforts.

## Data Availability

All data produced in the present work are contained in the manuscript.

## Funding

BMBF Verbundprojekt SARS-CoV-2Dx to Sebastian Drube

Leibniz Center for Photonics in Infection Research (LPI-BT6, CEZ), Germany’s Excellence Strategy (Balance of the Microverse, C.E.Z)

## Appendix

### Supplementary material 1

Search syntax Pubmed:

*(“COVID-19”[Mesh] OR “COVID-19”[tw] OR “COVID 19”[tw]) AND (“Treg*”[tw] OR “regulatory T*”[tw] OR “T regulatory cell*”[tw] OR “T-regulatory cell*”[tw] OR “foxp3*”[tw] OR “T-Lymphocytes, Regulatory”[Mesh]) NOT (Review[pt])*

Search syntax Web of Science:

*TS=(“COVID-19”) AND TS=(“Treg*” OR “regulatory T*” OR “T regulatory cell*” OR “T-regulatory cell*” OR “foxp3*”)*

### Supplementary material 2

PRISMA Checklist (see external file)

